# Reducing the Cost of Rapid Antigen Tests through Swab Pooling and Extraction in a Device

**DOI:** 10.1101/2021.03.11.21252969

**Authors:** Tim Berking, Sabrina G. Lorenz, Alexander Ulrich, Joachim Greiner, Clemens Richert

## Abstract

The COVID-19 pandemic places a significant stress on the viral testing capabilities of many countries. The value of rapid point-of-care (PoC) antigen tests is becoming increasingly clear, but implementing frequent large scale testing is costly. We have developed an inexpensive device for pooling swabs, extracting specimens, and detecting viral antigens with a commercial lateral flow assay detecting the nucleocapsid protein of SARS-CoV-2 as antigen. The holder of the device can be produced locally through 3D printing. The extraction and the elution can be performed with the entire set-up encapsulated in a transparent bag, minimizing the risk of infection for the operator. With 6 swabs holding approx. 0.1 mL specimen each and 0.35 mL extraction buffer, 43±6 % (n= 8) of the signal for an individual extraction of a positive control standard was obtained. Image analysis still showed a signal-to-noise ratio of ≥ 7 upon further eight-fold dilution. Our current total cost of materials is below $ 2 per tested person or 20% of our cost for an individual PoC test. These findings suggest that pooling can make frequent testing more affordable for schools, universities and other institutions, without decreasing sensitivity to an unacceptable level. Further validation of the method is required.

## Introduction

Identifying asymptomatic spreaders of viral infections is an important task in a pandemic. This is particularly true for COVID-19, a viral disease with many ‘silent spreaders’ that is proving difficult to control within a population.^1,2^ Frequent testing of a large portion of the population is expected to reduce the spread of the disease, because spreading events can be avoided. Testing is most effective when performed frequently, and by using an approach that provides results quickly after collecting the specimen, even if the method is less sensitive than RT-PCR.^3^ Lateral flow assays (LFAs) that detect a viral antigen are among the rapid tests that are attractive in this context.^4^ They produce results within 15-30 min without the need for a laboratory and can show excellent specificity.^5^ Several studies have validated lateral flow rapid antigen tests in the current pandemic.^6,7,8,9,10^

Even though lateral flow antigen tests can be mass produced, the cost of mass testing individuals on a regular basis may become prohibitive, even for developed countries.^11,12^ Pooling of samples that are then analyzed in a single procedure is known to lower the cost of testing in scenarios with low incidence.^13^ Pooling has been reported for PCR tests detecting SARS-CoV-2,^14,15,16^ and a recent report from a company suggests that pooled samples can successfully be analyzed by lateral flow point of care (PoC) test cassettes,^17^ but pooling has not become an established practice for rapid antigen tests. One reason for this may be the need for biosafety measures and laboratory equipment to perform the pooling steps, which counterbalances the ease of use for LFAs at the point of need. These complications may be overcome by an inexpensive device suitable for the pooling, extraction and analysis of specimens. Here we report such a device, together with results from exploratory pooling experiments in a laboratory setting.

## Materials and Methods

Components of the Panbio™ COVID-19 Ag Rapid Test Device (nasopharyngeal) were used (Abbott Rapid Diagnostics Jena GmbH, Jena, Germany), for which the manufacturer reports a sensitivity of 91.4% (nasopharyngeal swab versus nasopharyngeal PCR) and a specificity of 99.8% (https://www.globalpointofcare.abbott/en/product-details/panbio-covid-19-ag-antigen-test.html). The Abbott test detects the nucleocapsid protein of SARS-CoV-2 as antigen and contains a control line detecting chicken IgY. According to the manual, the extraction buffer of the test kit contains tricine as buffer component, NaCl, Tween 20 as non-ionic surfactant, sodium azide as bacteriostatic (listed as < 0.1 %), and Proclin 300 as other preservative/biocide. Dilution experiments with water used Kabi Ampuwa sterile water (Fresenius) for injection purposes. Positive control swabs provided with the Abbott kits were used as source of SARS-CoV-2 antigen. Either the Noble Biosciences NFS-1 nasopharyngeal swabs supplied with the Abbott test kit were used (‘50 µL swabs’) or singly wrapped, sterile, cotton wool swabs (5 mm head) with a wooden stem (150 mm length) from neoLab (Heidelberg, Germany) as larger specimen alternatives (‘100 µL swabs’). Nasopharyngeal or throat plus nose swab specimen were self-collected from healthy volunteers among the authors who had tested negative in professional tests performed by physicians using the SARS-CoV-2 rapid antigen test (Roche, Mannheim, Germany). Plastic bags were zip lock all-purpose/freeze bags made of polyethylene, holding up to 1 L volume, suitable for transporting liquids in carry-on luggage in the EU, or up to 3 L volume (both from QuickPack, Renningen, Germany). Syringes were 10 mL Amefa LUER single-use sterile syringes (B. Braun, Melsungen, Germany). The holder for the syringe body acting as pooling container and the lateral flow test cassette was produced by 3D printing on a Prusa MK2S printer with PLA (polylactide) as material, instructed by CAD data in STL format. Photographs were taken with the camera of a Samsung Galaxy S20+ (SM-G985F) cellular phone and image analysis was performed using the free program ImageJ (NIH). Details of the image analysis are provided in the Supplemental files.

## Results and Discussion

### Pooling Set-up

We defined the following criteria for the test method to be developed: The method had to use inexpensive materials, including sterile, singly wrapped swabs and a sterile mixing container. The analytical procedure had to be based on a commercial, well established LFA system, and all steps had to be feasible without additional biochemical components and without the need for laboratory equipment. These criteria led to the system described below.

Figure 1 shows two core components of our device. One of those components is a syringe for medical use that functions as container for the swabs and as vessel in which the mixing occurs. With the goal to achieve pooling for at least five specimens, we tested several syringe sizes and swabs and settled on 10 milliliter (mL) syringes with the central plunger removed as containers. They will hold approx. ten flocked nylon swabs of the type included in both the Abbott and the Roche rapid antigen tests and six conventional sterile flocked cotton swabs with wooden stem, without losing the mobility necessary for thorough mixing. The latter cost a fraction of the nylon swabs, hold twice as much specimen, and are more pleasant when applied in the mouth.

**Figure 1.**
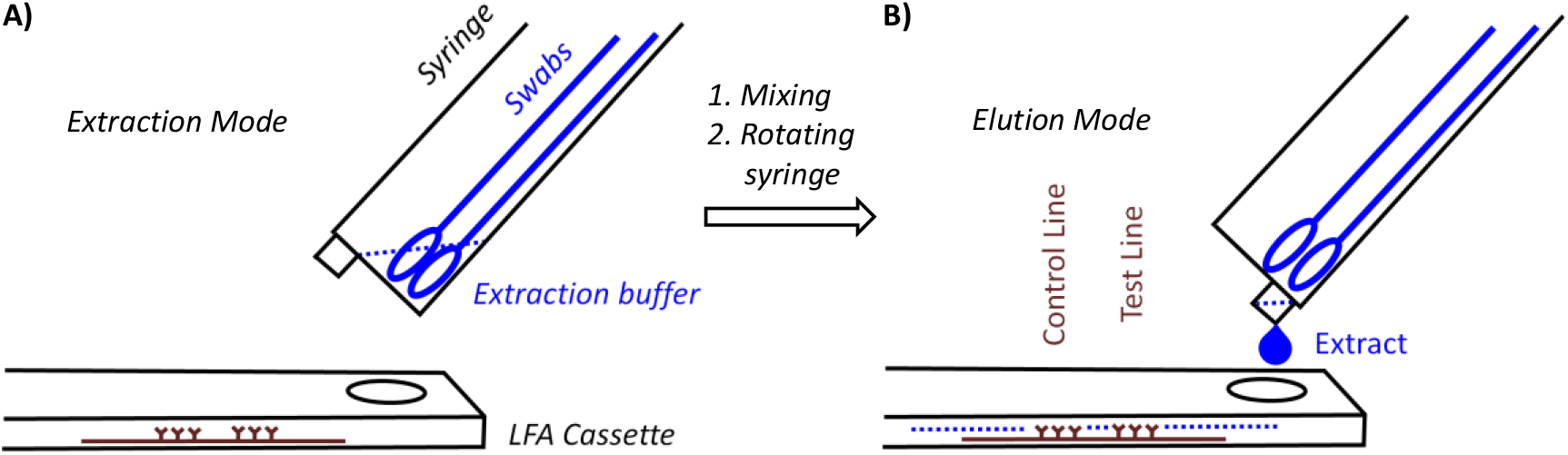
Principle of pooling and extraction in a syringe acting as container and elution onto a lateral flow assay (LFA) cassette for analysis. A) Container in extraction mode; thorough mixing is induced by rotating the bundle and moving individual swabs. B) Elution, induced by rotating the syringe by 180° to allow for the extract to drip onto specimen well of the LFA cassette.

The container is held at an angle to the LFA cassette. There are two positions for the syringe acting as container. In extraction mode, the LUER fit opening at the tip of the syringe is at its highest position to prevent premature leaking of the extract. The extraction buffer and the swabs are now entered, one after the other. Mixing is achieved by extensively rotating and moving the swabs immersed in the buffer for 3 minutes. This step requires some dexterity and experience to ensure that the liquid is homogeneous. It is easier to perform this step with more liquid, but dilution lowers the signal and is therefore undesirable. After mixing, the syringe is switched into elution mode, so that the LUER fit is in its lowest position, allowing the extract to drip into the specimen well of the cassette. To ensure that the necessary volume is applied, the swabs should be moved up and down to avoid blockage of the exit port, and the swabs should be gently squeezed against the wall of the syringe. Again, this calls for dexterity and may be rehearsed without potentially infectious material first by operators new to the device. It is also critical to perform the elution with close visual inspection to ensure that the necessary number of drops is applied.

Figure 2 shows the holder of the device and the final form of the syringe, as employed in our current procedure. The holder shown in Figure 2A has an arm with a circular opening for the syringe and indentations next to this opening that arrest the syringe in extraction or elution mode. The base of the holder has another indentation, where the LFA cassette is held in place. Further, there is a small indentation at the center bottom of the base plate, into which a stabilizing bar is pushed to provide mechanical stability to the assembly and to prevent it from tipping over during the extraction and elution steps.

**Figure 2.**
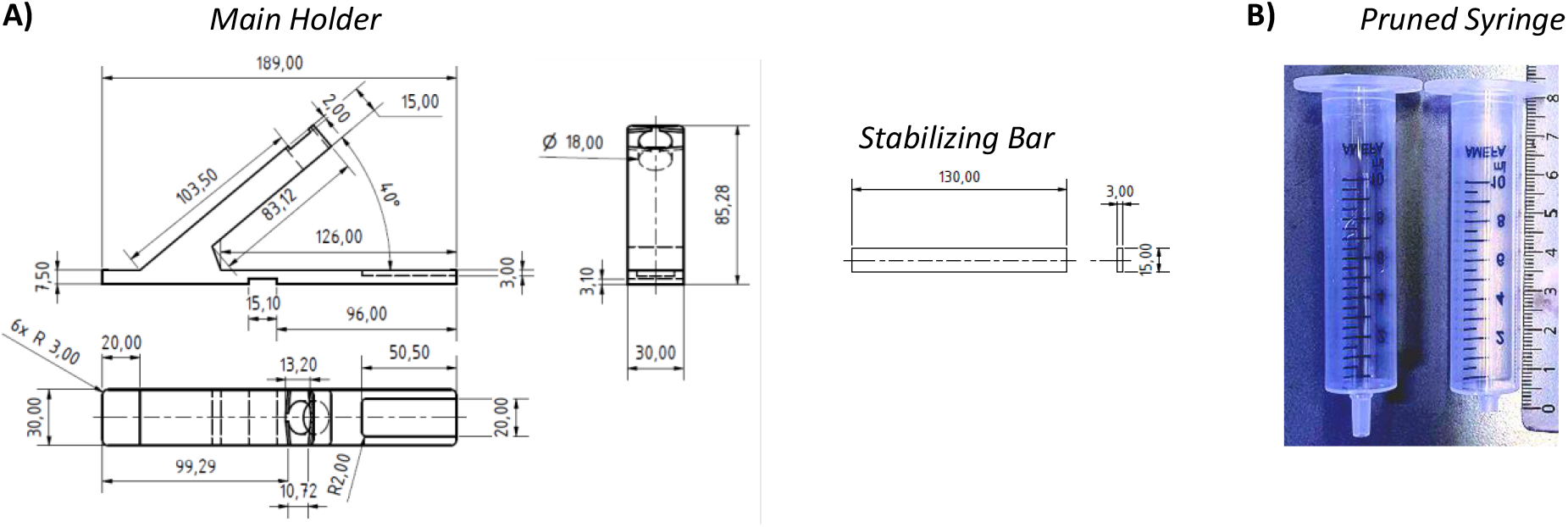
Components of the device. A) Technical drawings showing the dimensions (in mm) of the main holder, the stabilizing bar of the holder, and B) photograph of an unmodified 10 ml syringe next to a syringe used as pooling and extraction container that had its LUER fit pruned to reduce void volume. The length ruler next to the pruned syringe shows the length in cm.

While initial prototypes were made of solid PVC, both the main holder and the stabilizing bar are now produced by us by 3D printing in an inexpensive printer. In our currently preferred method for testing, the holder is disinfected and recycled when the test outcome is negative, but is disposed of when a test is positive. The cost of the material for printing both holder and stabilizing bar is less than $ 2, so that the method is inexpensive, even if the holder is treated as a disposable item in each test procedure with swab pooling.

One modification was made to the commercial 10 mL syringes used as container. Their LUER fit tip was pruned to avoid retention of a significant fraction of the extract in the form of the void volume of the device. Fully removing the tip led to uncontrolled elution and very large drops. Leaving the LUER opening unchanged retained too much of the valuable extract. The best performance was achieved with tips pruned to a length of 4 mm, as shown in Figure 2B. With this size opening, the volume of 100 µL, which is required for the lateral flow assay to function properly, elute in three large drops that can be visually observed, even if the entire set-up is encapsulated in a transparent polypropylene bag (Figure 3). We do the pruning of the tip with the blade of a carpet cutter, previously treated with disinfectant, while the syringe is in a steel holder (see Supplementary Information), but pruning may also be performed in a simpler, hand-held fashion.

**Figure 3.**
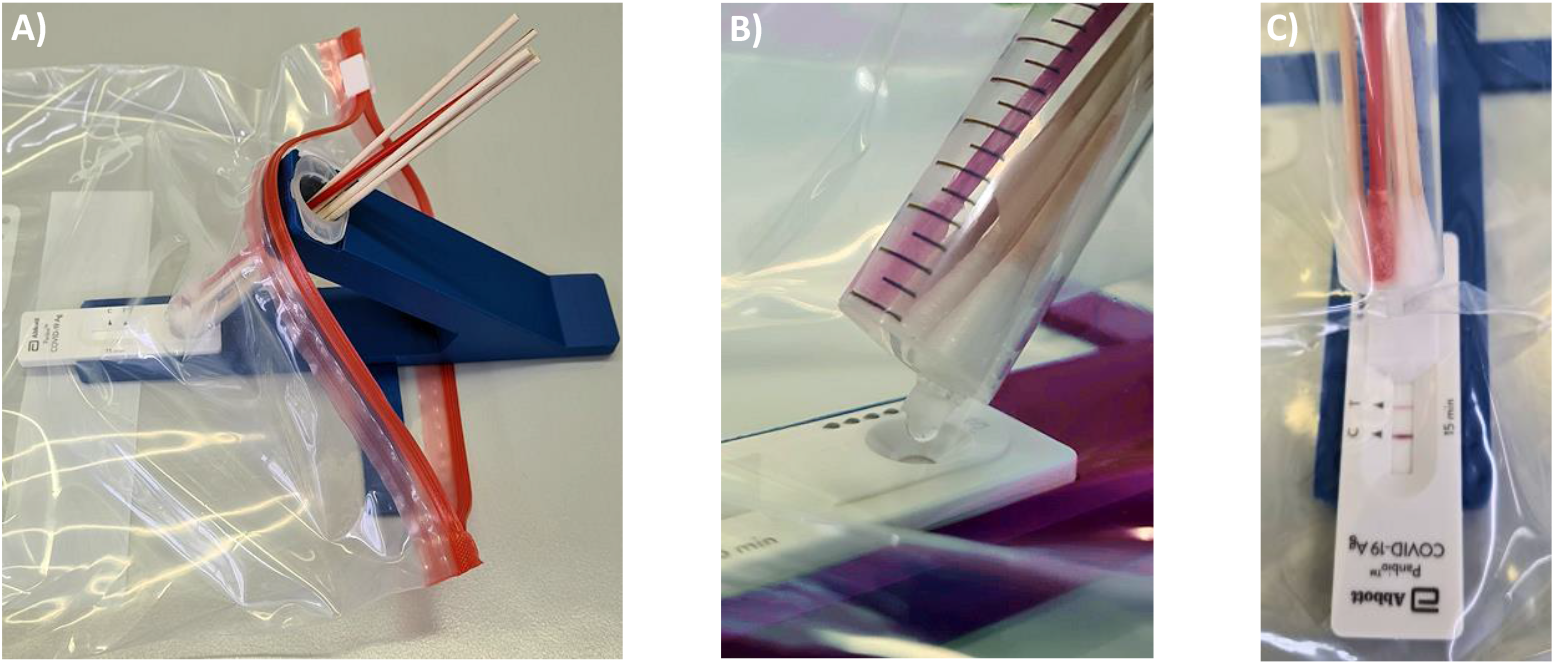
Photographs of testing with the device in a polyethylene bag with zip lock for encapsulation after loading. A) Device loaded with five 100 µL flocked cotton swabs holding specimen and one positive control swab (red). B) Close-up showing elution of the extract after thoroughly mixing for 3 minutes. Three drops are required to elute 100 µL from the pruned syringe, corresponding to 5 drops from the original Abbott extraction container. C) Result of a test, viewed through the encapsulating bag after 15 min.

Figure 3A shows how the entire set-up may be inserted into a sealable, transparent plastic bag of sufficient mechanical stability and optical transparency. Our preferred procedure involves self-sampling in a mixed saliva/nasopharyngeal mode under the supervision of a medically trained member of the team (‘superuser’) undergoing team testing. Sampling occurs in a fume hood or well ventilated area with a minimum distance of 3 m between individual team members and superuser. The superuser sets up the device, drips the necessary volume of extraction buffer into the syringe, places the set-up in the transparent bag and supervises the collection of the specimens. The members of the team enter their swabs into the container, one by one, wearing an inexpensive polyethylene glove on the hand with which they handle the swab, and the superuser, wearing medical-grade gloves and a surgical mask or FFP2 mask, then closes the bag and sprays it down with disinfectant from the outside. After a brief interval to allow for evaporation of the alcohol-based disinfectant, the superuser performs the extraction, elution and analysis while the entire set-up is inside the closed bag (Figures 3B/C).

### Extraction and Analytical Assay

Pooling is expected to increase the total volume of the extract. To minimize the resulting loss in signal, the concentration of the antigen should be kept as high as possible. To achieve this, the test procedure was adjusted. First, we gravimetrically determined the volume absorbed by swabs after immersion in water for 5 seconds, with the aid of an analytical balance. We found that the NFS-1 flocked style nylon swabs provided with the Abbott test kit absorb 56.7 ± 3.0 µL under our conditions. Given the uncertainty of the viscosity of the real specimen, we refer to these swabs as ‘50 µL swabs’, assuming that they will take up approx. 50 µL specimen when applied in saliva plus nasal sampling. The less expensive 5 mm cotton wool swabs with wooden stem took up 101.3 ± 4.1 µL of water under the same conditions, and we refer to those swabs as ‘100 µL swabs’.

We assumed that thorough mixing after pooling of the swabs produces a homogeneous liquid as extract. The total volume of this extract will be the sum of the volume of the extraction buffer employed and the volume entered with the specimens. Again, for simplicity, we assumed that the latter volume is either 50 µL or 100 µL per person participating in the team test. The standard protocol, as described in the user manual for the Abbott Panbio COVID-19 Ag Rapid Test calls for 300 µL extraction buffer. The bottle provided with the test kit (25 tests) contains 9 mL or 360 µL per test. Assuming that some residue will remain, we used up to 350 µL of this, which equals 13 drops from the Abbott bottle. Unless otherwise noted, one positive control swab was included in all assays as a reliable and constant source of antigen to be detected.

Figure 4 shows typical results from assays performed with our device, and Table 1 lists the results obtained. The numerical values for signal intensities of the test (SARS-CoV-2 antigen) and control lines were obtained by analyzing photographs taken with the camera of a smartphone and analyzed with the free software ImageJ, available from the National Institutes of Health, as detailed in the Supplementary Information. The plots below each photograph of the read-out window of the test cassettes in Figure 4 are the results of the integration of the gray value data calculated in ImageJ.

**Table 1.** Results of rapid antigen test with or without pooling of swabs.

| Entry No. | No. of swabs <sup>a)</sup> | Type of swabs used [$\mu\text{L}$ capacity] <sup>b)</sup> | Estimated $V_{\text{tot}}$ [$\mu\text{L}$] incl. specimen | Dilution with additional liquid | Relative intensity test/control [%] | Signal relative to simplex assay<sup>c)</sup> [%] |
| --- | --- | --- | --- | --- | --- | --- |
| 1 | 1 | 50 | 300 | - | 86 | 100 |
| 2 | 6 | 50 | 600 | - | 47 | 55 |
| 3 | 6 | 100 | 1100<sup>d)</sup> | wet swabs <sup>e)</sup> | 32 | 37 |
| 4 | 6 | 100 | 850 | - | $37 \pm 5$ <sup>f)</sup> | $43 \pm 6$ <sup>f)</sup> |
| 5 | 6<sup>g)</sup> | 100 | 950 | - | 0 | - |
| 6 | 6 | 100 | 850 | 2-fold <sup>h)</sup> | 24 | 28 |
| 7 | 6 | 100 | 850 | 4-fold <sup>h)</sup> | 13 | 15 |
| 8 | 6 | 100 | 850 | 8-fold <sup>h)</sup> | 7 | 8 |

<sup>a)</sup> Unless otherwise noted, one of the swabs was a positive control swab (Abbott).
<sup>b)</sup> Positive control swab was added in dry form.
<sup>c)</sup> Simplex assay is assay with a single swab, according to the manufacturer's manual of the LFA test.
<sup>d)</sup> 300 $\mu\text{L}$ extraction buffer plus 300 $\mu\text{L}$ sterile water.
<sup>e)</sup> Pre-hydrated with sterile water prior to sampling.
<sup>f)</sup> Mean ± one standard deviation from eight experiments.
<sup>g)</sup> No positive control swab included.
<sup>h)</sup> Extract prepared as for entry No. 4, but then diluted with extraction buffer.

**Figure 4.**
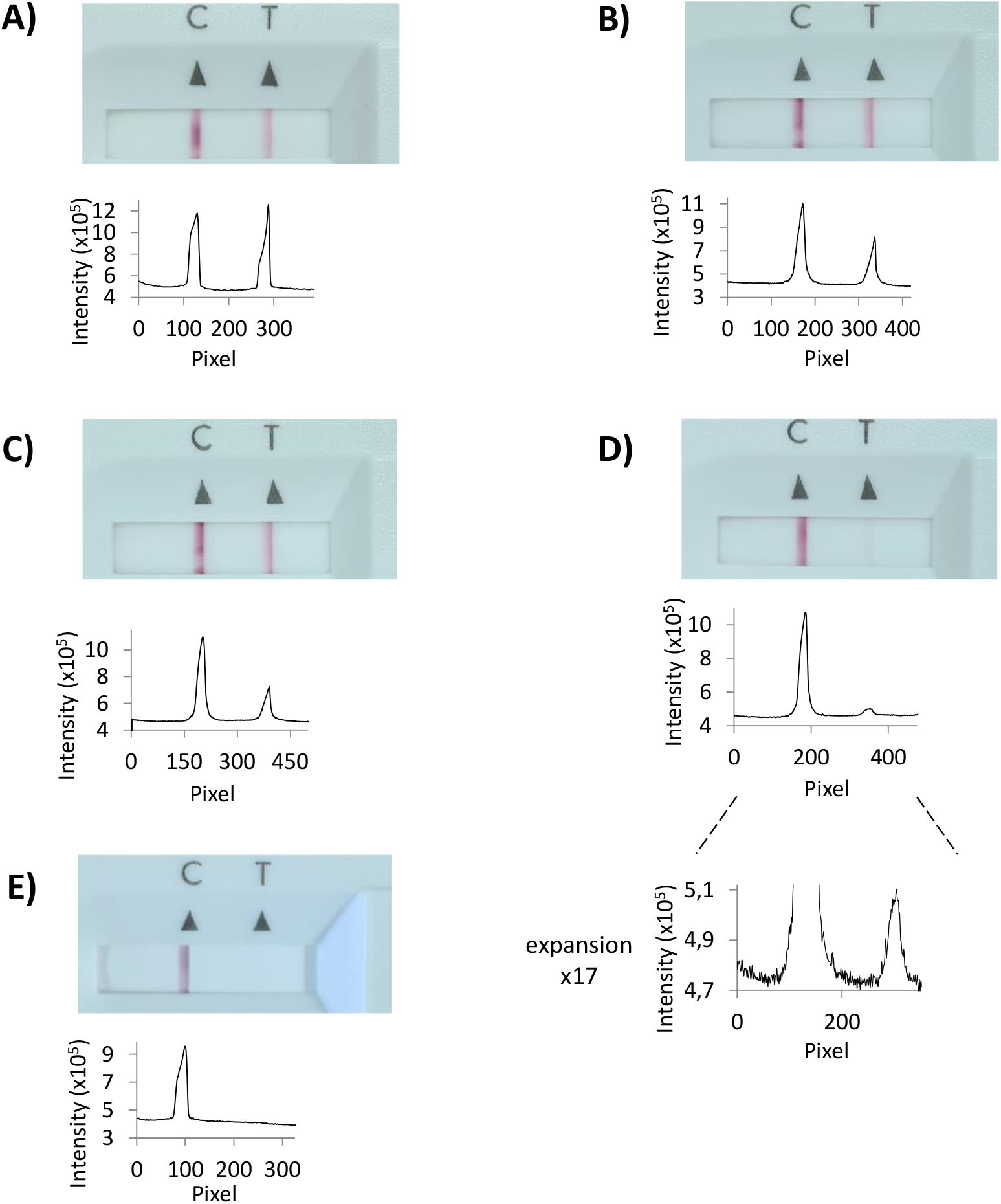
Representative results from rapid antigen tests with or without pooling. Photographs of the read-out window of lateral flow assay cassettes are shown, together with signal intensity plots for each test results, as obtained by integration with ImageJ. The entry number of Table 1 is listed for each graphic. A) Standard assay with a single positive control swab (entry 1). B) Pool of five nasopharyngeal 50 µL swabs with specimens of healthy volunteers and one positive control swab (entry 2). C) Five cotton wool 100 µL swabs and one positive control swab (entry 4). D) Same as C), but with 8-fold dilution with extraction buffer (entry 8). An expanded view of the integration plot is shown below the original plot. E) Six pooled cotton wool swabs from healthy volunteers (entry 5). All photographs were taken 15 min after applying the extract to the test cassette.

For the conventional, single-swab assay, run as recommended in the user manual of the Panbio Abbott test, the intensity of the test line was 86% of that of the control line (detection of chicken IgY), as shown in Figure 4A and entry 1 of Table 1. When five of the 50 µL Noble Biosciences NFS-1 nasopharyngeal swabs with specimens of healthy volunteers plus the dry positive control swab were used with 350 µL of the extraction buffer, a signal corresponding to 55% of that for the individual test was found, as expected based on an approximate dilution factor of two (entry 2 of Table 1). This is the most conventional form of pooling, using original swabs, as included by the supplier of the test kit (one per cassette). Unfortunately, such swabs were quoted to us by the importing company at a price exceeding $ 2 per swab, if ordered separately from the test kits, making this option unattractive from a cost perspective in the current pandemic market situation. Since we also wished to improve the ratio of specimen volume to extraction buffer volume, we opted for the 100 µL swabs.

In a first set of experiments, we used wet 100 µL swabs, pre-exposed to sterile water and an extraction buffer diluted two-fold with the same water. The approach gave 37% of the reference signal, i.e. considerably more than the 27% expected based on the estimated dilution factor, but was not pursued further over concerns that the dilution may change the characteristics of the extraction medium^10^ and reduce the viral load, and that the extra cost (and effort) to provide individual sterile vials with dilution water to each participant of the team test would become prohibitive. Still, it is an interesting finding, as it suggests stronger binding of the antigen-loaded gold nanoparticles to the surface immobilized antibodies in medium with lower ionic strength than standard medium. Our previous studies with DNA-coated gold nanoparticles on other surfaces^18,19^ did not point in this direction, which may be due to the different biopolymers involved in the molecular recognition events. We also note that the dependence of signal on concentration may not be linear due to multivalency effects,^20^ as the gold nanoparticles can be expected to display more than one ligand per particle.

Our preferred protocol used sampling in the pharyngeal region of the mouth, with sufficient wetting with saliva, as well as collection of throat specimens (≥30 seconds total), followed by nasopharyngeal sampling. This approach ensures thorough wetting and avoids that partially wetted swabs take up too much of the extract solution, complicating elution. It is similar to sampling traditionally performed for PCR tests, and is in agreement with the conclusions of a recent study enrolling 659 patients that employed the Panbio test and showed excellent sensitivity for the combination of saliva and nasal sampling.^21^ When combined with the 350 µL of extraction buffer, available from the test kit, the 100 µL extract required for the lateral flow assay was obtained without the need for any additional fluid. When the team test was performed eight times, we obtained a signal intensity of 43 ± 6 % (mean ± one SD) of that for the single swab standard procedure (entry No. 4, Table 1).

To further validate the method, we performed experiments with a negative control swab or six specimen-holding swabs from healthy, negatively tested volunteers, neither of which gave a detectable signal (entry 5 of Table 1). Finally, we performed exploratory experiments to determine the limit of detection with our method. For this, remaining extract from one pooling assay of entry 4 of Table 1, was diluted with extraction buffer in 1:2 dilution steps, to give 2-, 4-, and 8-fold lower antigen concentration (entries 6-8, Table 1). Even for the most dilute extract, after 8-fold dilution, a positive result was detectable, both visually, and by image analysis after photography (Figure 4D). The latter showed a signal-to-noise ratio of approx. 7 for the test result, versus background, indicating that at least another factor of two in dilution is achievable without losing the ability to make a clear positive/negative call. The signal intensity of entries 6-8 of Table 1 scales roughly linearly with dilution, again suggesting that our method is producing results undistorted by the change in protocol.

We note that the results of Table 1 should be regarded as the lower limit of the signal we expect for a positive specimen. The positive control swab from Abbott is designed for an assay with 50 µL swabs, but 100 µL swabs are employed in our preferred low-cost version of the test. So, while the total volume has gone up approx. 3-fold with 6-fold pooling, the amount of specimen is increasing approx. two-fold over that of the standard swab, resulting in a numerical decrease in concentration of antigens by a factor of 1.5 only.

Even though only fluids intended by the original procedure are being used, a validation of the pooling approach with clinical specimens and field study specimens is required. Assuming that this validation will confirm the results of our exploratory experiments, the team test approach may facilitate surveillance for viral infections at approx. five-fold reduced costs at the 6-fold pooling level. We experienced better compliance with a twice weekly voluntary test regime when testing was performed as a team test rather than as individual tests with the need for medical appointments. For ordinary team members, the time effort is well below 5 min per test, and the effort of the superuser is approx. 30 min per team test. The ability to run the tests at the workplace with low set-up costs and minimized infection risk due to encapsulation, as well as the self-sampling option, contribute to the attractiveness of the team test approach. Follow-up in positive cases should be done using more sensitive methods, such as RT-PCR, and negative results should not be the basis for medical decisions.

We note that our procedure deviates from that recommended by Abbott and that is has not been validated by clinical studies. Our goal was to detect infectious individuals early, using a process that is sustainable, both financially and in terms of the time effort involved. The manual of the Panbio test states that it has a detection limit of 2.5×10^1.8^ TCID_50_ of SARS-CoV-2 and a sensitivity of 94.1% ‘with samples of Ct values ≤33’. Data from an earlier study,^7^ as well as data from a recent field study^22^ both indicate that the Abbott Panbio rapid antigen test reliably detects ≥ 90% of individuals with a viral load corresponding to a ct value of ≥ 28-30. This is at the upper limit of what is believed to make a person infectious.^7,9,23^ Therefore, we have reason to believe that the drop in sensitivity observed for 6-fold pooling keeps the detection limit within the range that successfully identifies active spreaders of COVID-19. Team tests may provide more safety to families, work teams, musicians, and other groups.

## Supporting information

protocols and cost analysis

stp files for 3D printing

## Data Availability

All data is being made available in the manuscript and the Supplementary files.

## Conclusions

With the pooling strategy described here, a roughly five-fold reduction in material cost may be achieved, while the accompanying reduction in sensitivity is only approx. two-fold, depending on the type of swabs employed. This reduction in sensitivity corresponds to approx. one ct number (or ‘average ct’),^22^ and may be compensated by increasing the sensitivity via quantitative image analysis, using a conventional smart phone and free image analysis software. Even without such compensating measures, a reduction by only one ct number should not outweigh the gain in practicability, which results from a simple, rapid, point-of-need procedure that does not require medical personnel or laboratory infrastructure. Pooling not only reduces the cost, but also allows the testing of more people when the availability of LFA test cassettes is limiting. Our approach minimizes the risk of infection when supervised self-sampling at a safe distance is practiced, and when the analytical procedure is performed in a sealed bag. Independent of the details of the protocol, the inexpensive device described here may help to reduce the transmission of SARS-CoV-2 and other viruses that threaten the livelihood of many communities. Our method should be validated further.

## Supplementary Data

Supplementary data is available online (protocol for image analysis, cost analysis and data for 3D printing).

## Acknowledgments

The authors thank Mathis Knabe, Dr. Claudia Loser, Dr. Eric Kervio, Daniela Göhringer, Jennifer Bremer, Juri Eyberg, Tim Gniech and the members of the Richert Research Group for their help. This work was supported by intramural funds. The work of the Richert Research Group on antiviral agents is supported by Volkswagen Foundation (supplement to grant Az 92 768).

