## Supplementary material for "Reducing the Cost of Rapid Antigen Tests through Swab Pooling and Extraction in a Device": protocols and cost analysis

for manuscript entitled

by

Tim Berking, Sabrina G. Lorenz, Alexander Ulrich, Joachim Greiner and Clemens Richert

**Contents**

1. Protocol for Image Analysis
2. Process of pruning syringes
3. Cost calculation for materials used
4. 3D Printing files

### 1. Image Analysis

Pictures of test cassettes for the numerical analysis of signal intensities were taken with a Samsung Galaxy S20+ (SM-G985F) smartphone in photo mode, using the wide angle option, without a flashlight, in 3:4 format. The test cassettes were photographed in daylight/fluorescent bulb laboratory light with an attempt to achieve even light exposure on the lab bench where pictures were taken.

We also tested the pro mode (ISO 100, 1/90s shutter speed, auto focus, 4000-5000 K white balance) for raw picture data, and compared them to the data in photo mode to verify that the picture is not falsified by the AI of the smartphone used. Both data showed no discernable difference in relative signal intensity.

The pictures were transferred to a laptop *via* USB cable and opened in ImageJ (downloaded from the ImageJ homepage). In subsequent steps, the pictures were turned into a 8-bit type and inverted, as documented below.

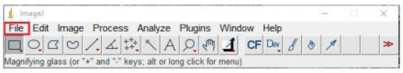


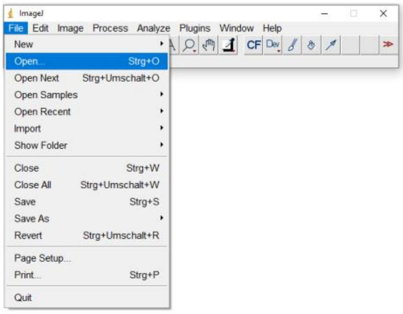


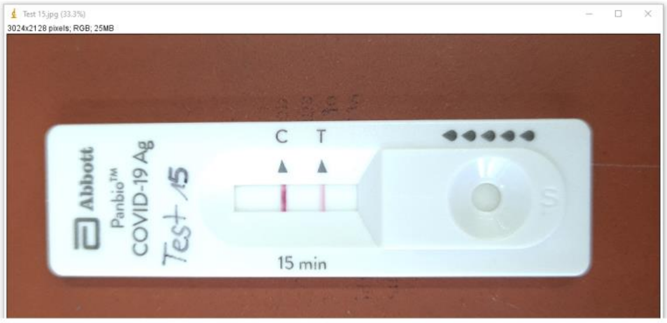


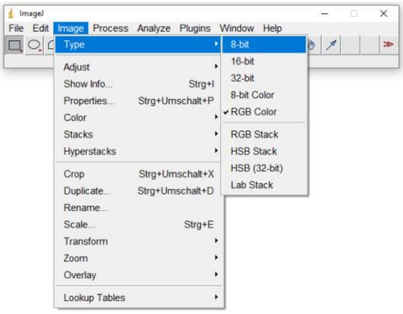


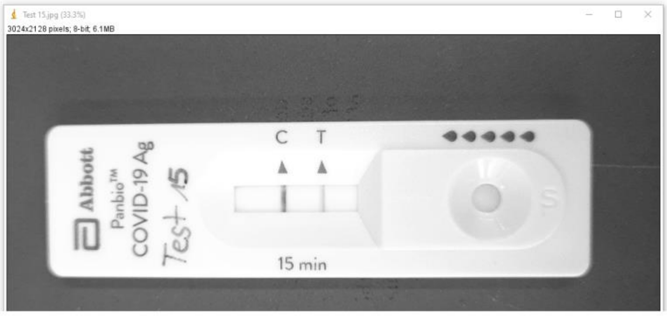


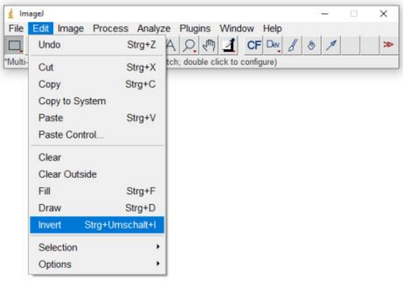


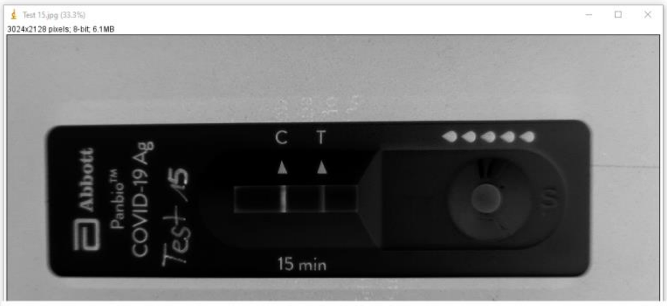


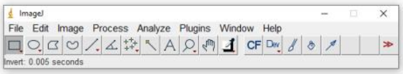

Rectangular areas of the identical size were chosen and marked around the control line, background (area between control line and test line) and the test line using “rectangle”. The gray value intensities were measured (Tab “Analyze” → Set Measurements), adjusted by activating "Integrated density" and automatically listed in the table “Results” as “IntDen”, as documented graphically below.

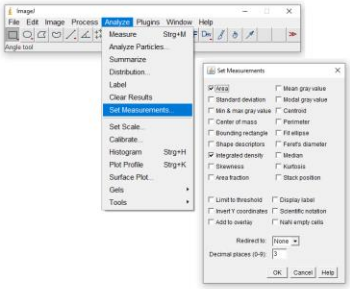

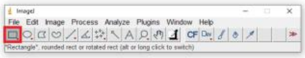

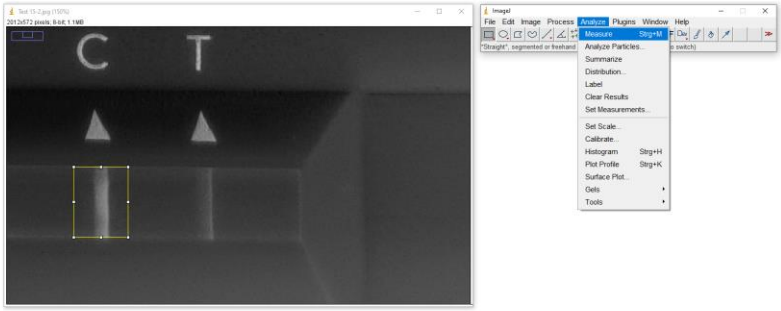

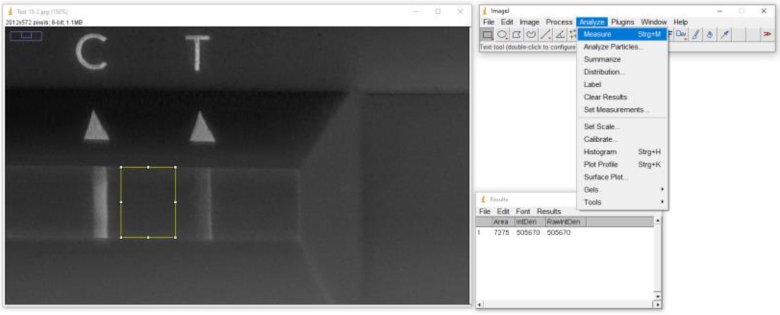

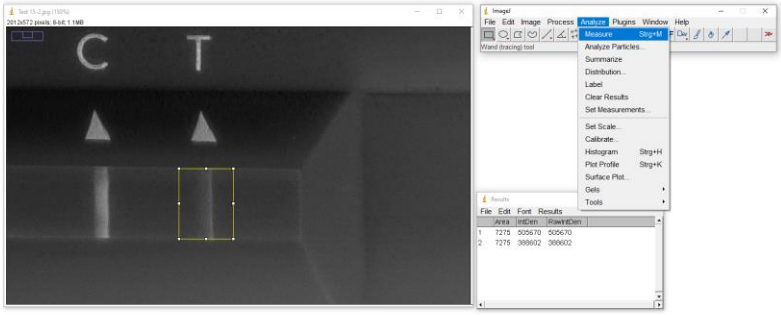

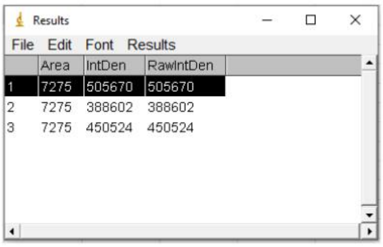

To calculate the relative signal intensity of test line versus control line, equation 1 was used:

$$Intensity\ test\ relative\ to\ control\ [\%] = \frac{intensity\ test\ line - intensity\ background}{intensity\ control\ line - intensity\ background} * 100 \qquad (1)$$

Furthermore, we used equation 2 to calculate the intensity of the test result of our pooling tests in relation to the simplex assay involving standard extraction of a single positive control swab (Abbott):

$$Intensity\ test\ related\ to\ simplex\ assay\ [\%] = \frac{Intensity\ test\ relative\ to\ control\ [\%]}{intensity\ test\ relative\ to\ control\ of\ simplex\ assay} \qquad (2)$$

#### 2. Pruning Syringes

The LUER outlet of the syringe should be shortened to avoid a loss of extract volume that otherwise remains in the opening.

***Components employed in the procedure***

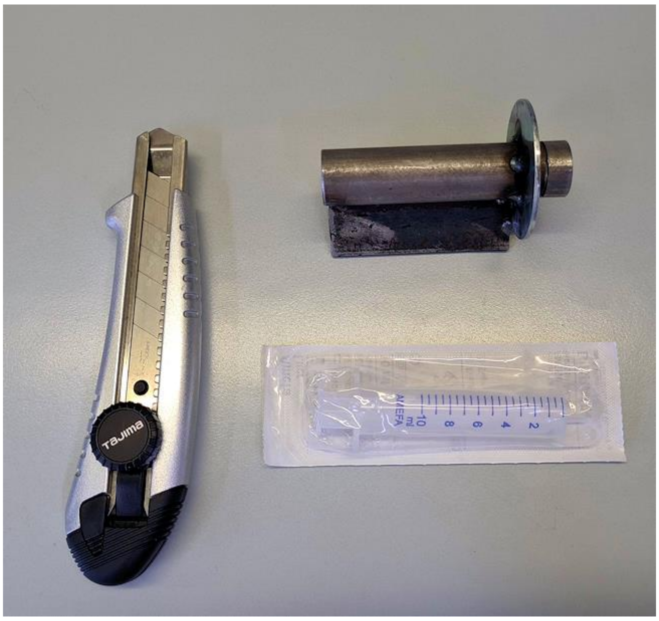


***Pruning the tip of the syringe in the holder***

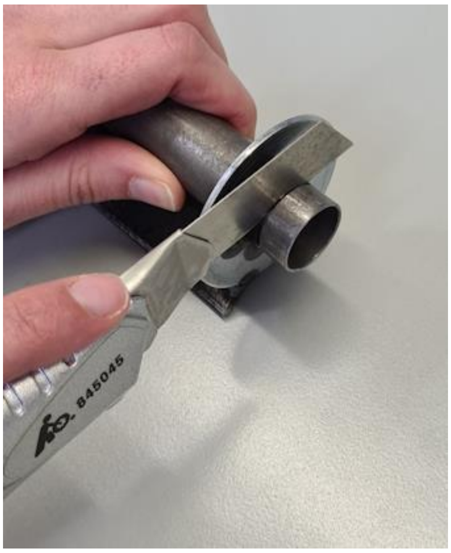

#### 3. Cost of Materials Employed

The following is a list of current costs to us in Euros (€), including local VAT (19%). Disinfectant and the face mask of the superuser are not included, assuming that they are standard equipment.

| **Material of Component** | **Supplier** | **Items per packing unit** | **Cost per packing unit** | **Cost per item** | **Items per test** | **Cost per test** |
|---|---|---|---|---|---|---|
| LFA components | Abbott | 25 | 193,38 | 7,74 | 1 | 7,74 |
| Holder and bar | IFB | 1 | 1,54 | 1,54 | 0,1[a] | 0,15 |
| Transparent bag | Aldi | 15 | 1,65 | 0,11 | 1 | 0,11 |
| Syringe | Amefa | 100 | 6,00 | 0,06 | 1 | 0,06 |
| MedComf gloves | Neolab | 100 | 1,70 | 0,02 | 5 | 0,09 |
| Medical gloves | Stockroom | 100 | 15,00 | 0,15 | 2 | 0,30 |
| Cotton wool swabs | Neolab | 100 | 12,50 | 0,13 | 6 | 0,75 |
| | | | | | Cost per Team Test (6-fold pooling) | 9,19 |
| | | | | | **Cost per person** | **1,53** |

[a] Assuming that one team test out of ten is positive, resulting in the disposal of the holder .

#### 4. 3D Printing files

The data for instructing a 3D printer (.stp format) is being made available as two separate files.
